# THE IMPACT OF LOCKDOWN ON PUBLIC HEALTH DURING THE FIRST WAVE OF COVID-19 PANDEMIC: LESSONS LEARNED FOR DESIGNING EFFECTIVE CONTAINMENT MEASURES TO COPE WITH SECOND WAVE

**DOI:** 10.1101/2020.10.22.20217695

**Authors:** Mario Coccia

## Abstract

What is hardly known in the studies of the COVID-19 global pandemic crisis is the impact of general lockdown during the first wave of COVID-19 pandemic both public health and economic system. The main goal of this study is a comparative analysis of some European countries with a longer and shorter period of national lockdown during the first wave of COVID-19 from March to August 2020. Findings suggests that: a) countries with shorter period of lockdown have a variation of confirmed cases/population (%) higher than countries with longer period of lockdown; b) countries with shorter period of lockdown have average fatality rate (5.45%) lower than countries with longer period of lockdown (12.70%), whereas variation of fatality rate from August to March 2020 suggests a higher reduction in countries with longer period of lockdown (−1.9% vs 0.72%). However, Independent Samples Test and the Mann-Whitney test reveal that the effectiveness of longer period of lockdown versus shorter one on public health is not significant. In addition, the COVID-19 pandemic associated with longer period of lockdown has a higher negative impact on economic growth with consequential social issues in countries. Results of the impact of COVID-19 lockdowns on public health and economies of some leading countries in Europe, during the first wave of the COVID-19 pandemic, can provide vital information to design effective containment strategies in future waves of this pandemic to minimize the negative effects in society.

## INTRODUCTION

The main goal of this study is to explain the effect of a policy response of lockdown on public health and economy to reduce the impact of severe acute respiratory syndrome coronavirus 2 (SARS-CoV-2), which is the strain of novel coronavirus that causes Coronavirus disease 2019 (COVID-19) in society. This study focuses on data of the first wave of COVID-19 pandemic (from March to August, 2020) in countries that have applied longer or shorter period of lockdown to assess the effective reduction of infected cases and fatality rates as well as the impact on Gross Domestic Product. Lessons learned from this study can be important to design effective public responses for constraining future waves of the COVID-19 and similar epidemics of new infectious diseases.

In the presence of COVID-19 pandemic, and in general of vast epidemics, as governments have to cope with consequential threats on public health, it is important, very important to analyze what containment measures are effective and which are not. Nicoll and Coulombier (2009, p. 3ff) argue that containment measures have the goal to stop as many transmissions of infectious disease as possible. In particular, governments take actions to constrain/prevent chains of transmission and outbreaks, through vigorous contact tracing, quarantine of contacts, general lockdown of people and economic activities, etc. The crux of the study here is rooted in the concept of *lockdown* as policy response of countries to cope with diffusion of pandemic in society and some brief backgrounds are useful to understand and clarify it. The dictionary by Merriam-Webster (2020) defines lockdown as: “a temporary condition imposed by governmental authorities (as during the outbreak of an epidemic disease) in which people are required to stay in their homes and refrain from or limit activities outside the home involving public contact (such as dining out or attending large gatherings)”. Atalan (2020) shows that COVID-19 pandemic can be contained with a containment measures of social restrictions, such as lockdown. Tobías (2020, p. 2) states that: “Lockdown, including restricted social contact and keeping open only those businesses essential to the country’s supply chains, has had a beneficial effect”. This containment measure, in the presence of a pandemic or epidemic, has a variable period and includes one or more of actions, such as: school and workplace closing, cancellation of public/private events and closure of museums, restrictions on mass gatherings in public and private places, stay at home requirements, restriction on internal mobility and international travel, etc. (Nicoll and Coulombier, 2009; Petherick et al., 2020). Atalan (2020) argues that countries can decide to start the lockdown when there is an acceleration of daily confirmed cases beyond a critical threshold and to end it when there is a strong reduction of Intensive Care Unit (ICU) admissions. In general, lockdown as policy responses has main effects on public health, environment and economies of countries (Chakraborty and Mait, 2020).

### What is already known on these topics is based on manifold studies

Islam et al. (2020) argue that early application of the control measure of lockdown can generate a reduction of the incidence of COVID-19. The model by Balmford et al. (2020) also reveals that countries that applied immediately lockdown reduced deaths compared to countries that delayed the application of this strong containment measure. Chaudhry et al. (2020) explain, analyzing 50 countries having high number of confirmed cases of COVID-19, that 40 countries applied a full lockdown, 5 a partial one and 5 curfew only with different effects. In addition, this study suggests that lockdowns and pervasiveness of testing in society were not associated with COVID-19 mortality per million people, but full lockdowns and decreased country vulnerability to biological threats were significantly associated with the increase of patient recovery rates (Chaudhry et al., 2020). Gatto et al. (2020), at March 2020, analyzing results of their transmission model, argue that restriction to mobility and human interactions can reduce transmission dynamics of the COVID-19 by about 45%. Tobías (2020) shows that after the first lockdown in Spain and Italy, the slopes of daily confirmed cases, of deaths and of Intensive Care Unit (ICU) admissions have been flattened, but the natural dynamics of COVID-19 pandemic has not changed the underlying trend that is continued to increase. However, the second lockdown, based on more containment measures for mobility, seems to have changed the trend, reducing daily diagnosed cases, total deaths and ICU admissions. Other studies show the effects of COVID-19 lockdown on environment and in particular on level of air pollution. Briz-Redón et al. (2021) analyze changes in air pollution during COVID-19 lockdown in Spanish cities and show that lockdown has reduced the atmospheric levels of NO_2_, CO, SO_2_ and PM_10_, except the level of O_3_. Ghahremanloo et al. (2021) analyze the impact of COVID-19 containment measures on air pollution levels in East Asia and confirm that the concentrations of pollutants in February 2020 are lower than those of February 2019. In addition, NO_2_ also had significant reductions in Beijing-Tianjin-Hebei regions, Wuhan, Seoul, and Tokyo. In this context, Liu et al. (2021) analyze the effects of COVID-19 lockdown in about 600 major cities worldwide and show that NO_2_ air quality index value falls more precipitously relative to the pre-lockdown period, followed by PM_10_, SO_2_, PM_2.5_, and CO, but the level of O_3_ increases. Moreover, Liu et al. (2021) argue that the impact of COVID-19 lockdown on environmental pollution generates health benefits in terms of the expected averted premature deaths due to air pollution declines. In general, the evidence that COVID-19 lockdowns have reduced levels of air pollution and detrimental effects of polluted environment on human health is now rarely contested. However, *what is hardly known in these research topics* is whether and how the application of general lockdown during the first wave of COVID-19 has been or has not been effective to reduce the negative impact on public health and economic system. This investigation is part of a large research project on factors determining the transmission dynamics of the COVID-19 pandemic and its socioeconomic impact. Results of the study here can explain the effects of lockdown in the first wave of COVID-19 in society and can be important, very important to design effective strategies and support sustainable technologies to cope with future waves of COVID-19 and future epidemics of new infectious diseases^1^.

## DATA AND STUDY DESIGN

### 1.1 Data and their sources

The study here focuses on six countries in Europe having a comparable institutional and socioeconomic framework: three countries with a shorter period of lockdown and three with a longer period of lockdown. In particular:

□ *Countries with a shorter period of lockdown* are (average about 15 days of lockdown):
  – Austria from 3/16/2020 to 4/13/2020, 29 days
  – Portugal from 3/19/2020 to 4/2/2020, 15 days
  – Sweden did not apply any lockdown
□ *Countries with a longer period of lockdown* are (average roughly 61 days of lockdown):
  – France from 3/17/2020 to 5/11/2020, 56 days
  – Italy from 3/09/2020 to 5/18/2020, 71 days
  – Spain from 3/14/2020 to 5/09/2020, 57 days
□ Period under study: from March to August 2020, indicating the first wave of the COVID-19 pandemic.

The study here considers data of confirmed cases, fatality rate and GDP aggregates in these countries after the application of lockdown, i.e., from 15 April to 30 August 2020, a period that indicates the first wave of COVID-19 pandemic. These data provide important information to assess the effectiveness of policy responses based on lockdown to cope with COVID-19 global pandemic crisis. Data about public health are from Johns Hopkins Center for System Science and Engineering (2020) and economic data are from Eurostat (2020).

### 1.2 Measures

▪ *Numbers of COVID-19 related infected individuals* are measured with confirmed cases of COVID-19 divided by population % of countries under study
▪ *Numbers of COVID-19 related deaths* are measured by fatality rate of COVID-19 given by total infected individuals divided by deaths (%) of countries
▪ *Economic activity of countries* is measured with Gross Domestic Product (GDP) and main components (output, expenditure and income). Unit of measure is chain linked volumes, index 2010=100. The accounting period is the calendar quarter (Q), based on 2019-Q2, 2020-Q1 and 2020-Q2 (Q1= January, February, March; Q2=April, May, June). Quarterly national accounts data are a vital instrument to economic analysis and policy and in assessing the state of the business cycle (cf., Coccia, 2010).

### 1.3 Data analysis procedure

*Firstly*, data are analyzed with descriptive statistics, using a comparative approach between countries with longer and shorter period of lockdown, considering arithmetic mean of confirmed cases standardized with population, of fatality rates from April to August 2020 and of the quarterly national accounts of GDP. In addition, to assess the effects of lockdown on public health, it is calculated the average variation of confirmed cases standardized with population and fatality rate from 15 April 2020 to 30 August 2020, a period indicating the first wave of the COVID-19 pandemic.

*Secondly*, in order to assess whether the difference of arithmetic mean and average variation of confirmed cases standardized with population, fatality rate and GDP aggregate between countries with shorter and longer period of lockdown is significant, the Independent Samples *t*-Test is performed. In particular, the Independent Samples *t*-Test compares the means of two independent groups in order to determine whether there is statistical evidence that the associated population means are significantly different. The null hypothesis (*H*_0_) and alternative hypothesis (*H*_1_) of the Independent Samples *t*-Test are:

*H*_0_: µ_1_ = µ_2_, the two population means are equal in countries with shorter and longer period of lockdown

*H*_1_: µ_1_ ≠ µ_2_, the two population means are not equal in countries with shorter and longer period of lockdown Considering the small sample, the nonparametric Mann-Whitney *U* Test is also performed to compare whether there is a difference in the dependent variable for these two independent groups. It compares whether the distribution of the dependent variable (i.e., confirmed cases standardized with population and fatality rate) is the same for the two groups and therefore from the same population.

*Thirdly*, the study represents the trends of average value of infected individuals and fatality rates of countries understudy from April to August 2020 aggregated in two groups, given by:

□ Countries with a shorter period of lockdown are (average about 15 days)
□ Countries with a longer period of lockdown are (average roughly 61 days)

The study analyzes these trends with simple regression model, using the specification of a linear relationship:

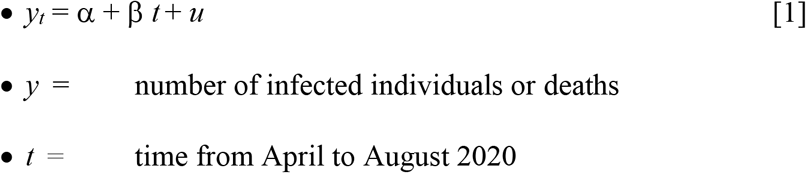

Ordinary Least Squares (OLS) method is applied for estimating the unknown parameters of linear models [1]. Statistical analyses are performed with the Statistics Software SPSS® version 26.

## RESULTS

### □ Impact of COVID-19 and lockdown on public health

Figure 1 reveals that countries with shorter periods of lockdown have a lower average values of confirmed cases/population (%) but a higher variation of confirmed cases/population (%) than countries with longer periods of lockdown form April to August 2020 (the first wave of the COVID-19 pandemic).

**Figure 1.**
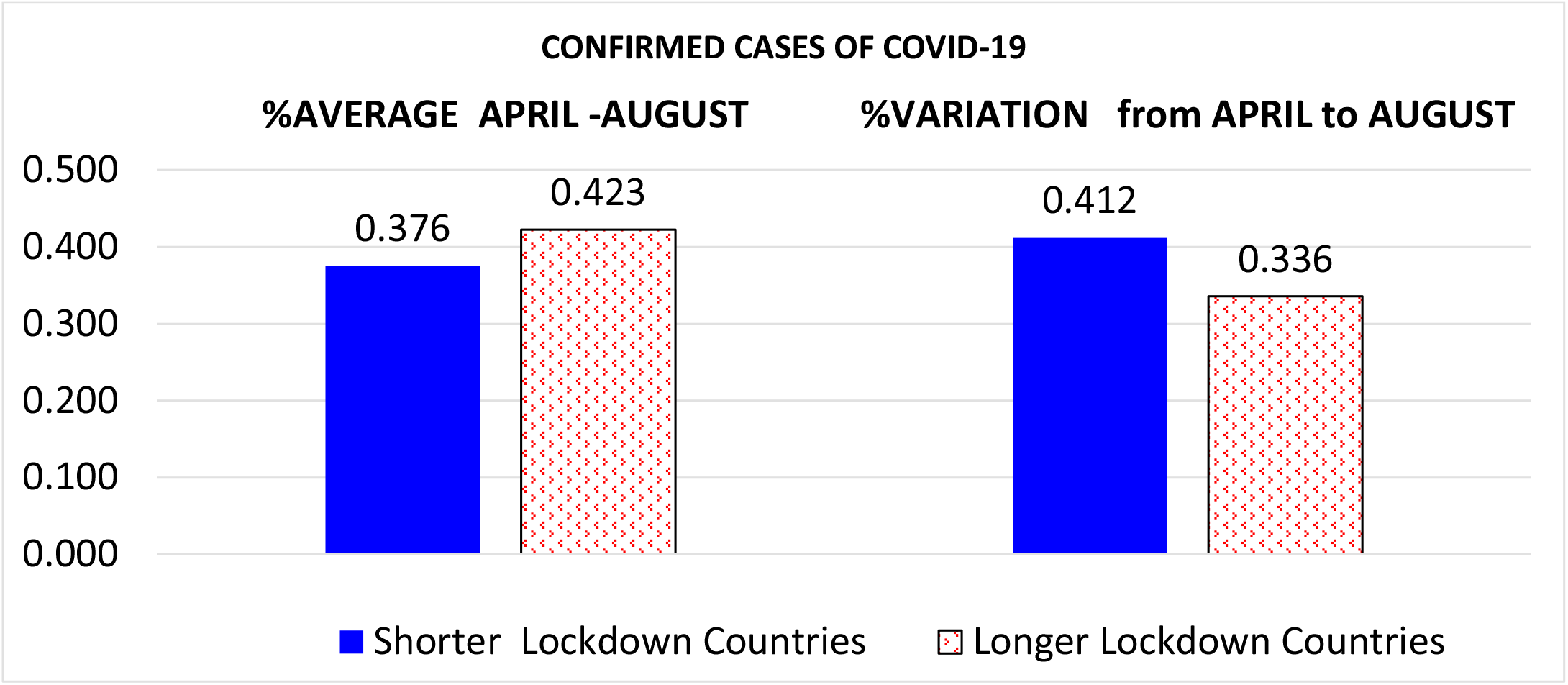
Average values and average variation of confirmed cases/population (%) over April-August 2020 in countries with shorter and longer period of lockdown.

Figure 2 reveals that countries with shorter periods of lockdown have a lower average magnitude of fatality rates (%) and a reduction of fatality rate lower than countries with longer period of lockdown over April - August 2020 (−0.72% vs. −1.90%). In order to assess the significance of the difference of arithmetic mean and average variation of confirmed cases standardized with population and fatality rates between countries with shorter and longer period of lockdown, the Independent Samples *t* Test is performed; considering the small sample under study also the nonparametric Mann-Whitney *U* Test is performed as countercheck to reinforce results.

**Figure 2.**
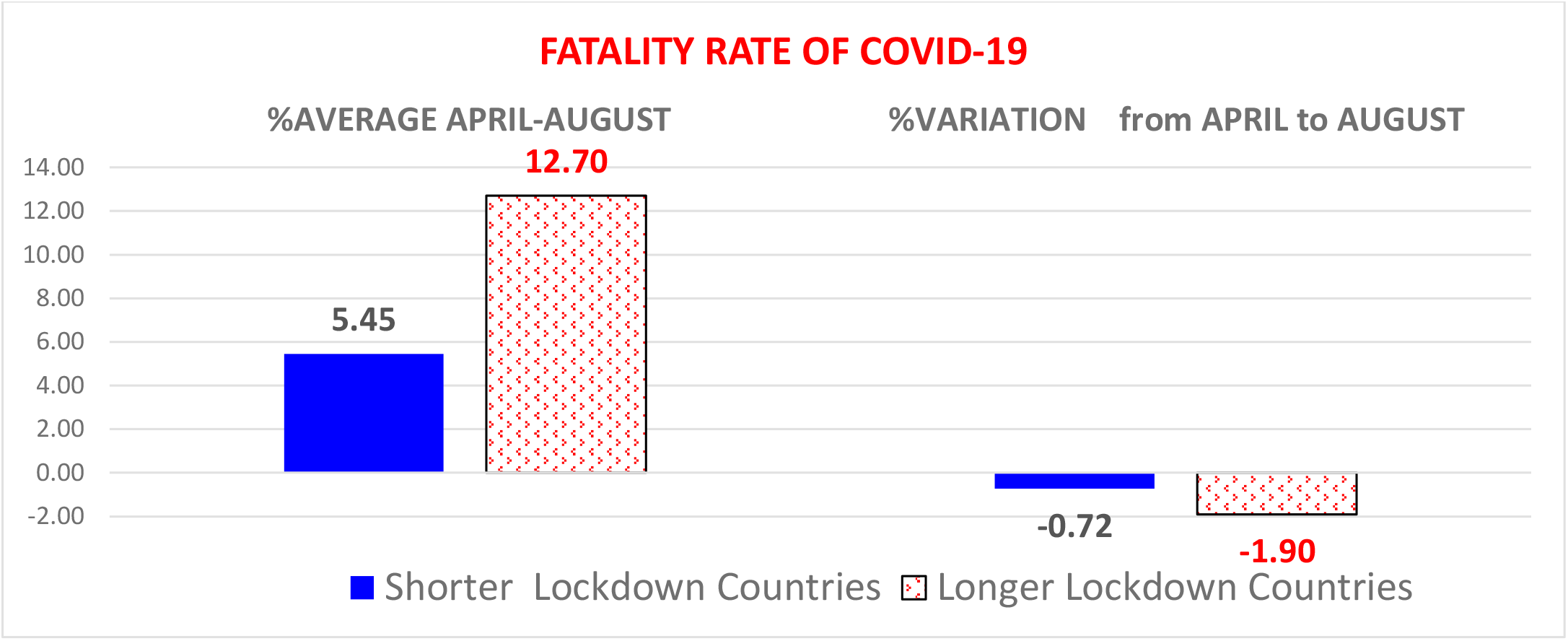
Average values and average variation of fatality rate (%) over April-August 2020 in countries with shorter and longer period of lockdown.

The *p*-value of Levene’s test is not significant, and we have to consider the output of “Equal variances assumed”. Based on results, there is a significant difference in mean days of lockdown (*t*_4_ = −4.825, *p* < .01) and average fatality rates (*t*_4_ = −3.343, *p* < .05) between countries with longer and shorter days of lockdown. In particular, the average fatality rate of countries with shorter period of lockdown was −7.3 percent points lower than countries with longer period lockdown because of higher initial incidence. Other indicators are not significant.

Tables 3 and 4, based on Mann-Whitney test, show that fatality rate in the group with shorter period of lockdown is significantly lower than the group of countries having a longer period of lockdown (*U* = 0, *p*-value = .005). Other indicators also here are not significant.

**Table 1.** Descriptive statistics for the impact of lockdown on public health, period April-August 2020.

| Period April-August 2020 | groups | N | Mean | Std. Deviation | Std. Error Mean |
| --- | --- | --- | --- | --- | --- |
| Days of lockdown | 1 | 3 | 14.670 | 14.503 | 8.373 |
|  | 2 | 3 | 61.330 | 8.386 | 4.842 |
| Average cases/population | 1 | 3 | 0.004 | 0.002 | 0.001 |
|  | 2 | 3 | 0.004 | 0.001 | 0.001 |
| Average fatality rate | 1 | 3 | 0.055 | 0.032 | 0.018 |
|  | 2 | 3 | 0.127 | 0.020 | 0.012 |
| Variation average cases/population | 1 | 3 | 0.004 | 0.003 | 0.002 |
|  | 2 | 3 | 0.003 | 0.002 | 0.001 |
| Variation fatality rate | 1 | 3 | -0.007 | 0.012 | 0.007 |
|  | 2 | 3 | -0.019 | 0.020 | 0.011 |

*Note:*
group 1= countries with a shorter period of lockdown (Austria, Portugal, Sweden)
group 2= countries with a longer period of lockdown (France, Italy and Spain)

**Table 2.**
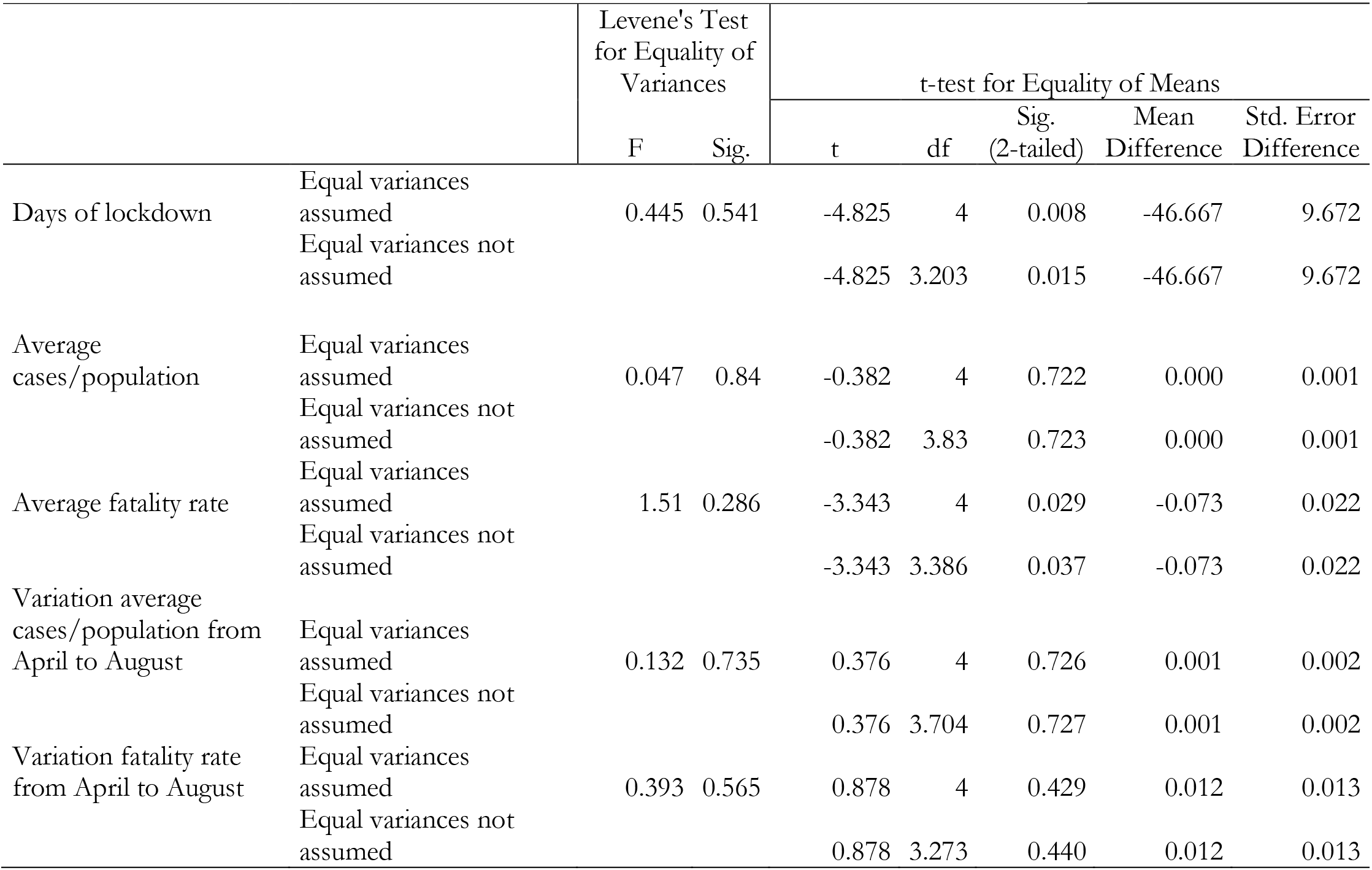
Independent Samples Test for the impact of lockdown on public health.

**Table 3.** Mann-Whitney Test. Rank for the impact of lockdown on public health.

| Period from April to August, 2020 | Groups | N | Mean Rank | Sum of Ranks |
| --- | --- | --- | --- | --- |
| Days of lockdown | 1 | 3 | 2 | 6 |
|  | 2 | 3 | 5 | 15 |
|  | Total | 6 |  |  |
| Average cases/population | 1 | 3 | 3 | 9 |
|  | 2 | 3 | 4 | 12 |
|  | Total | 6 |  |  |
| Average fatality rates | 1 | 3 | 2 | 6 |
|  | 2 | 3 | 5 | 15 |
|  | Total | 6 |  |  |
| Variation average cases/population | 1 | 3 | 3.67 | 11 |
|  | 2 | 3 | 3.33 | 10 |
|  | Total | 6 |  |  |
| Variation fatality rate | 1 | 3 | 3.67 | 11 |
|  | 2 | 3 | 3.33 | 10 |
|  | Total | 6 |  |  |

*Note:*

**Table 4.** Mann-Whitney Test for the impact of lockdown on public health.

|  | Test Statistics<sup>a)</sup> |  |  |  |  |
| --- | --- | --- | --- | --- | --- |
|  | Days of lockdown | Average cases/population | Average fatality rates | Variation average cases/population from April to August | Variation fatality rate from April to August |
| Mann-Whitney U | 0 | 3 | 0 | 4 | 4 |
| Wilcoxon W | 6 | 9 | 6 | 10 | 10 |
| Z | -1.964 | -0.655 | -1.964 | -0.218 | -0.218 |
| Asymp. Sig. (2-tailed) | 0.05 | 0.513 | 0.05 | 0.827 | 0.827 |
| Exact Sig. [2*(1-tailed Sig.)] | .100<sup>b)</sup> | .700<sup>b)</sup> | .100<sup>b)</sup> | 1.000<sup>b)</sup> | 1.000<sup>b)</sup> |

Note: a) Grouping Variable: groups; b) Not corrected for ties.

Finally, table 5 does not provide significant results of estimated relationships maybe due to small sample. Figure 3 provides trends of confirmed cases and fatality rates that approximatively do not suggest a difference in the temporal dynamics of evolution of the COVID-19 pandemic in countries with longer or shorter period of lockdown. In particular, the reduction of fatality rates over time in groups under study here seems to be due to the favorable climate conditions of summer season that studies show how it can reduce the diffusion of the COVID-19 rather than different strategies of longer or shorter period of lockdown (cf., studies by Coccia, 2020, 2020a, 2020b, 2020c, 2020d; Rosario Denes et al., 2020).

**Table 5.** Estimated relationships, based on linear model of regression.

|  | *Confirmed cases*<br>*of shorter lockdown countries* | *Confirmed cases*<br>*of longer lockdown countries* | *Fatality rates*<br>*of shorter lockdown countries* | *Fatality rates*<br>*of longer lockdown countries* |
| --- | --- | --- | --- | --- |
| Constant $\alpha$ (St. Err.) | -5.34*** (.18) | -2.97***(.18) | 26.00*(8.88) | 21.95(14.06) |
| Coefficient $\beta$ (St. Err.) | 3.87E-10<sup>a</sup>(.00) | 2.156E-10<sup>a</sup>(.00) | -1.88E-9<sup>a</sup>(.00) | -1.58E-9<sup>a</sup>(.00) |
| Stand. Coefficient Beta | .995 | .896 | -.72 | -.48 |
| R<sup>2</sup> (St. Err. of Estimate) | .99(.00) | .77(.00) | .52 (.007) | .23 (.012) |
| F | 869.52*** | 34.42*** | 8.54* | 2.41 |

Note: a) not indicated; Dependent variable: *time*.
Significance:
\*\*\* $p$ -value<0.001
\*\* $p$ -value<0.01
\* $p$ -value<0.05

**Figure 3.**
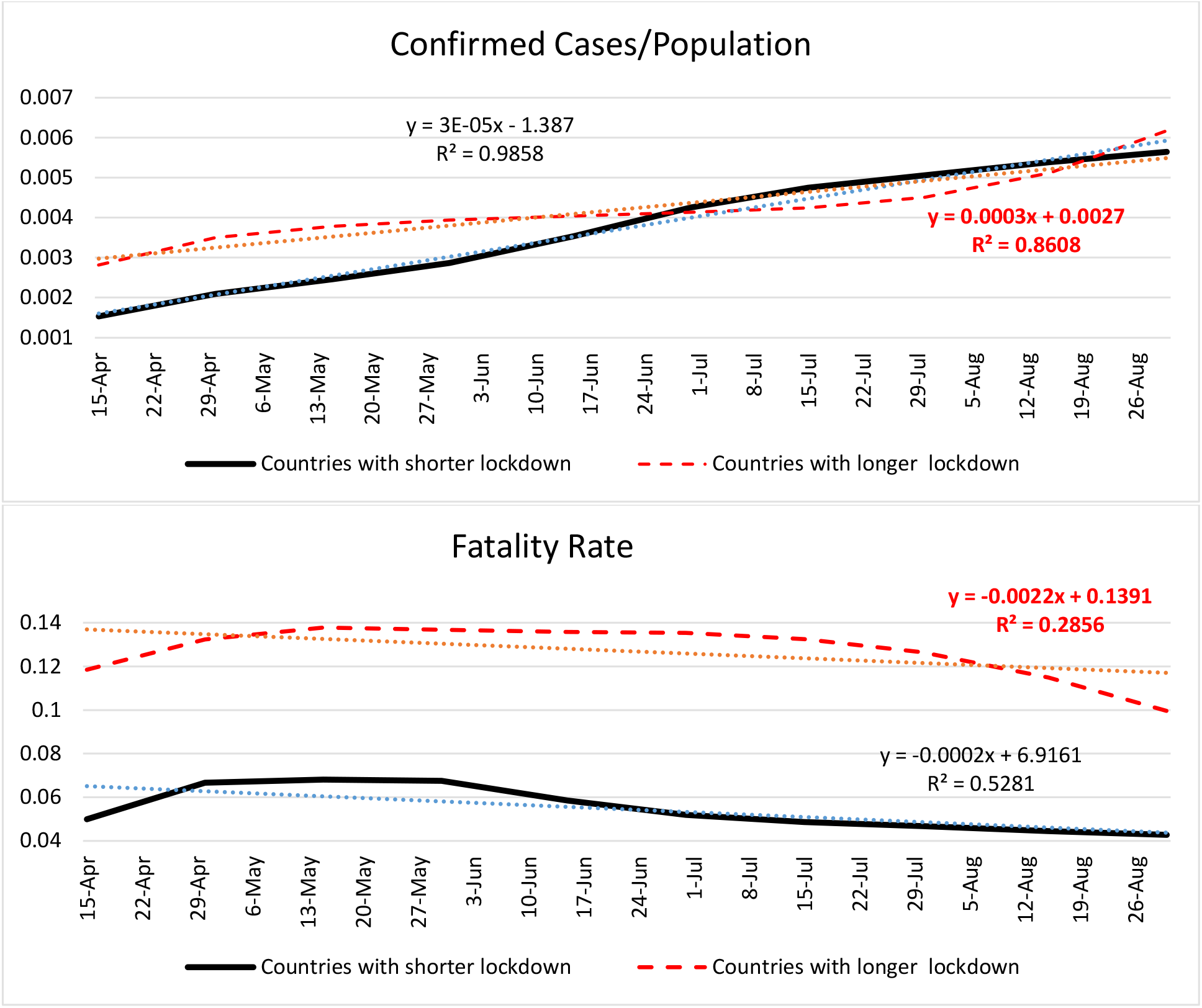
Trend of confirmed cases/population and fatality rates over April-August 2020 in countries with shorter and longer period of lockdown.

### □ Impact of COVID-19 and lockdown on economic system

Figure 4 and table 6 show *ictu oculi* that countries applying a longer period of lockdown they have had a higher reduction of GDP comparing the index of GDP of the second quarter 2020 to the same indicator in the same period of 2019 and comparing GDP of the second quarter 2020 to the first quarter (Q) of 2020.

**Table 6.** Group statistics for GDP aggregates.

|  | Countries | N | Mean | Std. Deviation |
| --- | --- | --- | --- | --- |
| $\Delta$ GDP(2020Q2-2019Q2) | Shorter period of Lockdown | 3 | -14.33 | 4.59 |
|  | Longer period of Lockdown | 3 | -21.37 | 2.76 |
| $\Delta$ GDP(2020Q2-2020Q1) | Shorter period of Lockdown | 3 | -9.33 | 4.37 |
|  | Longer period of Lockdown | 3 | -12.97 | 2.83 |

*Note:* Q=Quarter of the Gross Domestic Product, GDP; Q1= January, February, March; Q2=April, May, June

**Figure 4.**
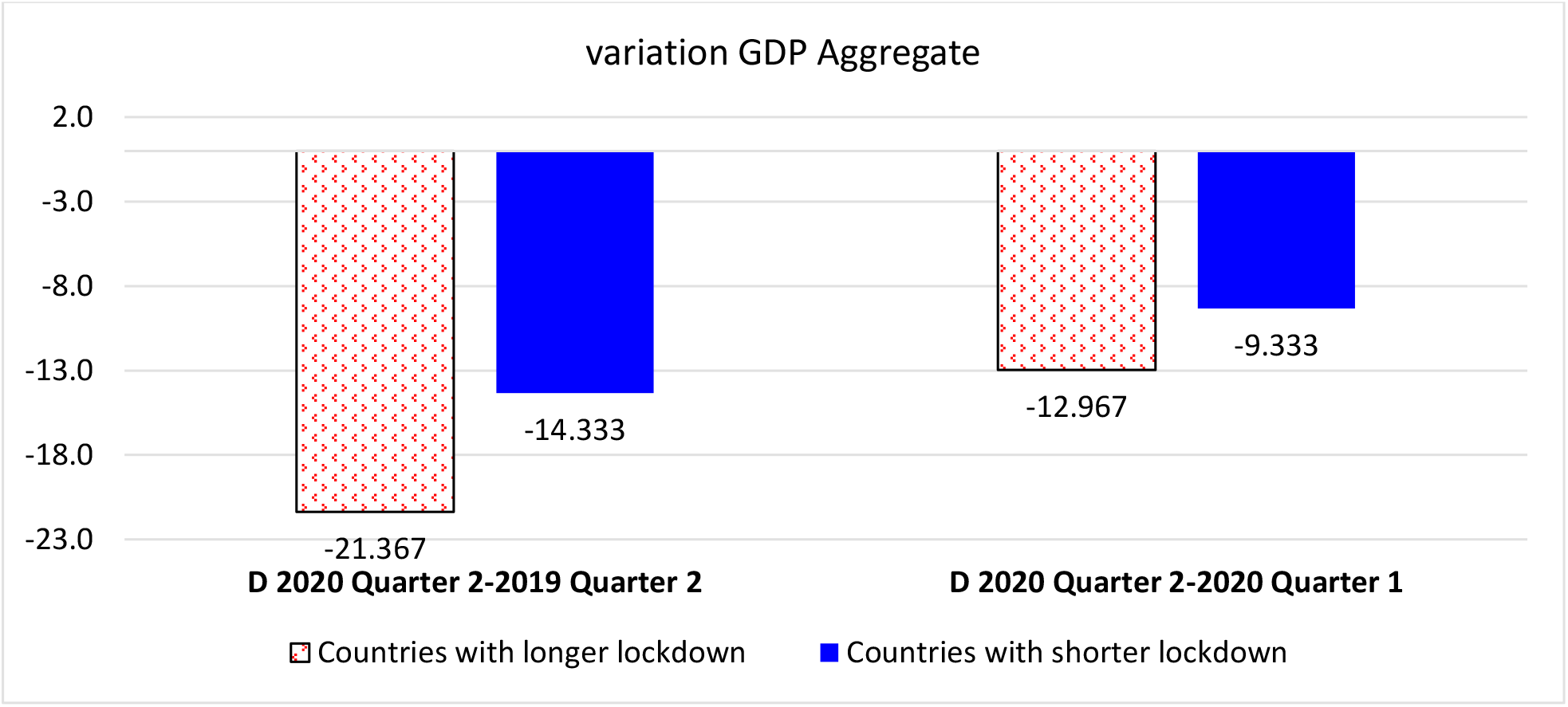
Variation of GDP aggregates (index 2010=100) from 2^nd^ quarter 2020 to 2^nd^ quarter of 2019 and from 1^st^ quarter 2020 to 2^nd^ quarter of 2020 between countries with longer and shorter period of lockdown. *Note*: Q1= January, February, March; Q2=April, May, June

Table 7 shows that the *p*-value of Levene’s test is not significant, and we have to consider the output of “Equal variances assumed”. Based on results, there is a significant difference in mean of GDP from Q2 in 2019 to Q2 in 2020 days between countries with longer and shorter period of lockdown (*t*_4_ = -2-274, *p* < .085). In particular, considering that these countries are in the same geoeconomic area, the GDP aggregate (index 2010=100) of countries applying a longer period of lockdown was about 7 points lower than countries applying a shorter period lockdown, likely due to systematic factor of deterioration of economic system given by the negative impact of COVID-19 pandemic and also different containment measures that have worsened this structural indicator of economic systems mainly in countries with longer periods of lockdown (cf., also Coccia, 2016).

**Table 7.**
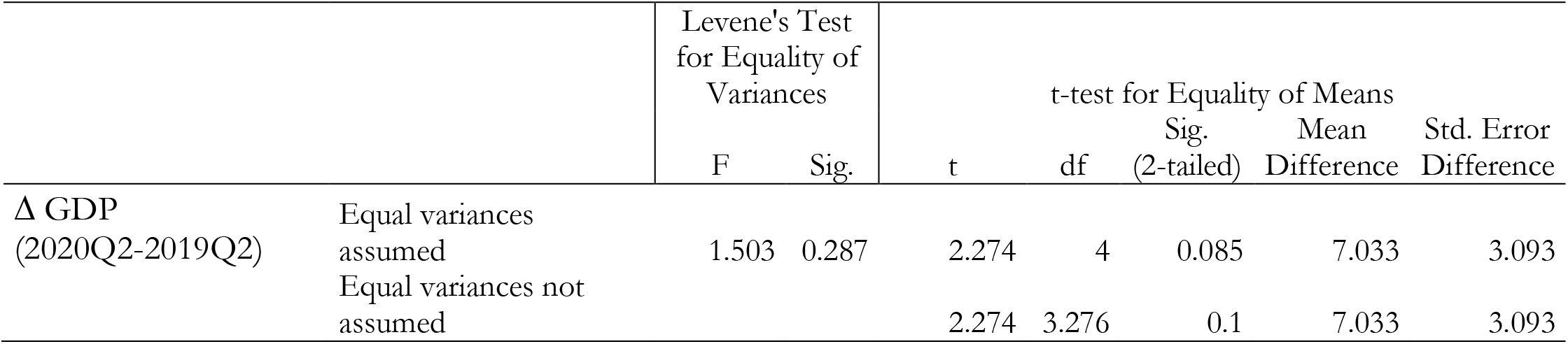
Independent Samples Test for the impact of lockdown on economy of countries.

|  |  | Levene's Test<br>for Equality of<br>Variances |  | t-test for Equality of Means |  |  |  |  |
| --- | --- | --- | --- | --- | --- | --- | --- | --- |
|  |  | F | Sig. | t | df | Sig.<br>(2-tailed) | Mean<br>Difference | Std. Error<br>Difference |
| $\Delta$ GDP<br>(2020Q2-2019Q2) | Equal variances<br>assumed | 1.503 | 0.287 | 2.274 | 4 | 0.085 | 7.033 | 3.093 |
|  | Equal variances not<br>assumed |  |  | 2.274 | 3.276 | 0.1 | 7.033 | 3.093 |

## DISCUSSION ON WHAT THIS STUDY ADDS

This study analyzes how different policy responses to COVID-19 based on longer or shorter periods of lockdown have affected public health and economic system. Previous studies suggest that measures of containment can constraint the human-to-human transmission dynamics of infectious diseases in different ways (Atalan, 2020; Prem et al., 2020; Tobías, 2020). However, to our knowledge, none investigations have performed a comparative analysis of the effects of longer or shorter period of lockdown on public health and economy of countries. *What this study adds to current studies* on the COVID-19 global pandemic crisis is that an accurate comparison of different government responses based on longer/shorter period of lockdown (from April to August 2020) to constraint the diffusion of COVID-19 pandemic, it suggests that a longer period of lockdown seems not to be associated with a statistically significant reduction of infected cases on population and variation of fatality rate, whereas countries applying a longer period of lockdown have a significant negative impact on economic system (given by contraction of real GDP growth % in 2020). In general, the COVID-19 pandemic tends to have natural dynamics that policy responses of lockdown at nation level seem to have a low impact in term of significant reduction of infected cases and mortality rates, but containment measures can slow down economic systems with consequent social issues. More specifically, results of the study here can be schematically summarized in the figure 5.

**Figure 5.**
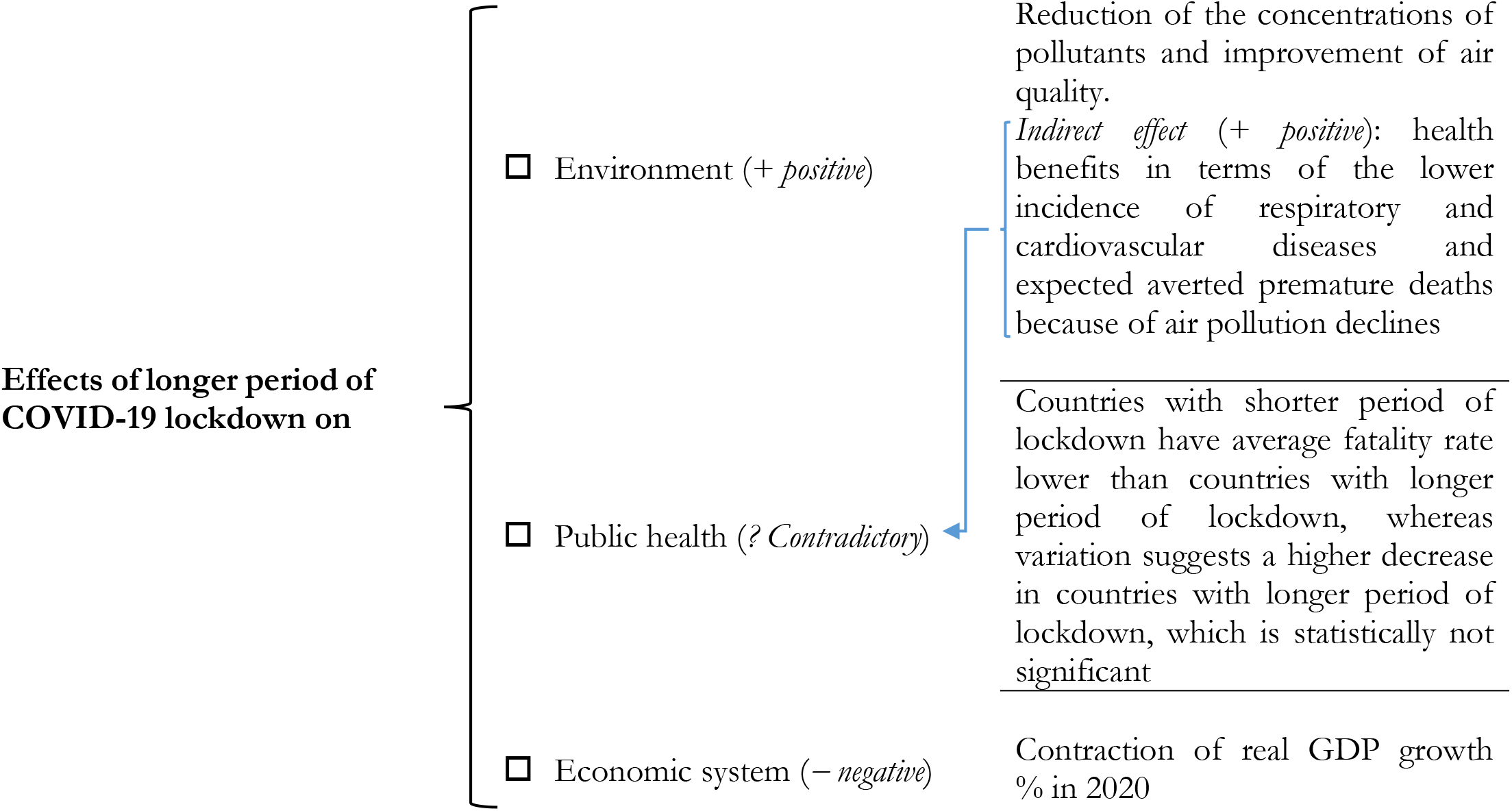
Impact of national COVID-19 lockdown on environment, public health and economies.

In general, the policy response of lockdown has the main aim, as containment measure, to reduce the impact of infectious disease on public health, but results here suggest contradictory and not significant effects on reduction of confirmed cases and fatality rates of longer period of lockdown compared to shorter period. However, longer period of lockdown has a main indirect positive effect on public health because of the reduction of concentrations of pollutants that improves air quality, lowering future incidence of respiratory and cardiovascular diseases and expected averted premature deaths because of air pollution declines (cf., Coccia, 2020, 2020d; Pope, 1989, 1996). In fact, Cui et al. (2020), based on a case study in China, show that reductions in ambient air pollution have avoided premature deaths and related morbidity cases, with main economic benefits in terms of reduction of public health expenses and improvement of social wellbeing.

Overall, then, countries with the on-going COVID-19 pandemic have showed an uncertain governance and an unrealistic optimism about their low vulnerability that a second wave of this pandemic cannot hit them (cf., Weinstein, 1987). In fact, although the severe impact on public health of the first wave of the COVID-19 pandemic, countries have shown still a low level of national planning to manage the second wave of the COVID-19 pandemic crisis with ambiguous and uncertain policy responses based on lockdown and other containment measures. In general, it seems that they have not used in comprehensive way the process of learning of the first wave of COVID-19 pandemic to cope with similar problematic situations, supporting effective and especially timely critical decisions (cf., Coccia, 2020; 2020e).

## CONCLUSIONS

The positive side of this study is that considers countries located in the same geo-economic area of the European Union having a similar social and democratic structure to perform a comparative analysis of containment measures to cope with COVID-19 pandemic. However, these results are based on a small sample of countries and future studies, to reinforce the generalization of these main findings, have to enlarge the sample, maintaining a comparable framework for statistical analyses. The statistical evidence here seems in general to show that the effects of longer *national lockdown* on public health in the first wave of COVID-19 pandemic are contradictory and not univocal, that is longer periods of lockdown seem not to significantly reduce confirmed cases and fatality rates, whereas can damage mechanisms of socioeconomic systems supporting the economic growth (cf., Coccia, 2019).

These conclusions are of course tentative because in the presence of the second and future waves of the COVID-19 pandemic and similar infectious diseases, manifold factors play a critical role. Countries are applying different policy responses of lockdown with different social restrictions in the presence of higher numbers of COVID-19 related infected individuals and deaths. However, the containment measure of lockdown, based on gradual and intermittent compulsory social restrictions, generates uncertain effects on the evolution of pandemic, public health and economic system.

Overall, then, there is need for much more detailed research on how countries in different economic, social, and institutional contexts can handle the COVID-19 pandemic crisis with different containment measures based on longer/shorter period of lockdown (Coccia, 2020e). To conclude, the investigation and explanation of the effects of shorter/longer period of lockdown on public health and economy are important, very important in order to design effective containment measures directed to minimize and/or contain the impact of second and third waves of COVID-19 outbreaks and future epidemics similar to the COVID-19 in society, as well as to not deteriorate the economic system of nations.

## Data Availability

The availability of all data is indicated in references.

https://ec.europa.eu/eurostat/databrowser/view/namq_10_gdp/default/table?lang=en

https://gisanddata.maps.arcgis.com/apps/opsdashboard/index.html#/bda7594740fd40299423467b48e9ecf6

## Footnotes

1 For studies about the interaction between science, technology and innovation for supporting socioeconomic systems, see: Coccia, 2017, 2017a, b; 2018, 2018a; Coccia and Wang, 2016; Coccia and Watts, 2020.

